# Pediatric COVID-19 in Southern California: clinical features and viral genetic diversity

**DOI:** 10.1101/2020.05.28.20104539

**Authors:** U Pandey, R Yee, M Precit, M Bootwalla, A Ryutov, L Shen, DT Maglinte, D Ostrow, JA Biegel, AR Judkins, JM Bender, X Gai, J Dien Bard

## Abstract

As the pandemic enters its fifth month, information regarding COVID-19 in children is rapidly evolving. Here, we explore clinical features and SARS-CoV-2 genetic variation in children presenting with COVID-19. We observed diverse clinical presentations and identified association between disease severity, viral load and age. SARS-CoV-2 genomes from the patients showed limited number of variations and an evolutionary rate comparable to other RNA viruses. We did not identify correlation between disease severity and viral genetic variations. Epidemiological investigation revealed multiple introductions of virus into Southern California.

## Introduction

Severe acute respiratory syndrome coronavirus–2 (SARS-CoV-2), the causative agent of coronavirus disease 2019 (COVID-19), was first reported in December 2019 in Wuhan, Hubei Province, China^1^. As of May 2020, there are over 1.7 million COVID-19 cases in the United States (US); however, information regarding COVID-19 in children is minimal. Although initially reported to have milder manifestation of COVID-19 compared to adults, studies have now linked COVID-19 to multisystem inflammatory syndrome (MIS-C) and gastrointestinal symptoms in children^2,3^. During a pandemic, the viral genome can be interrogated to understand the extent of the outbreak and to provide valuable insights into viral evolution and potential links to disease severity^4,5^. To date, California has over 99,000 confirmed COVID-cases and over 3,800 deaths^6^, among which vast majority of hospitalized patients are in Southern California. Currently, a single SARS-CoV-2 genome isolated in Southern California has been deposited in GISAID database, signifying paucity of genomics data on SARS-CoV-2 isolates circulating in Southern California^7^. In this study, we examined the clinical presentation of COVID-19 in children and assessed the genomic variations and epidemiology of the viral isolates using whole-genome sequencing (WGS) directly from clinical specimens.

## Results and Discussion

During an 8-week period we identified 35 pediatric patients with confirmed COVID-19, of which 22 (62.9 %) were seen at outpatient clinics and 13 (37.1%) were admitted to the hospital upon presenting to the emergency department. Demographics and clinical presentation are summarized in Table 1. The median age of the 35 patients was 12.5 years (range: 18 days to 18.5 years) with a male predominance (20/35, 57.1%). Median time to discharge of hospitalized patients was 4.0 days. While most reports suggest that SARS-CoV-2 causes asymptomatic to mild infections in children making them an important link in community-based viral transmission^8^, over half of our cohort was symptomatic. Among 20 patients with available medical history, 14 (66.7%) were symptomatic with the common symptoms being fever (57 %), congestion (36%), cough (36%), and shortness of breath (29%). Other observed symptoms included wheezing, chest pain, rhinorrhea, diarrhea, sore throat, and headache. Three of 4 patients (75%) with chest-imaging showed opacities in the lungs. Five patients (14%) required oxygen supplementation, of which 3 (60%) had a chronic condition. No patients reported travel history, however, four had direct contact to individuals with COVID-19. No death was observed in our cohort.

**Table 1.**
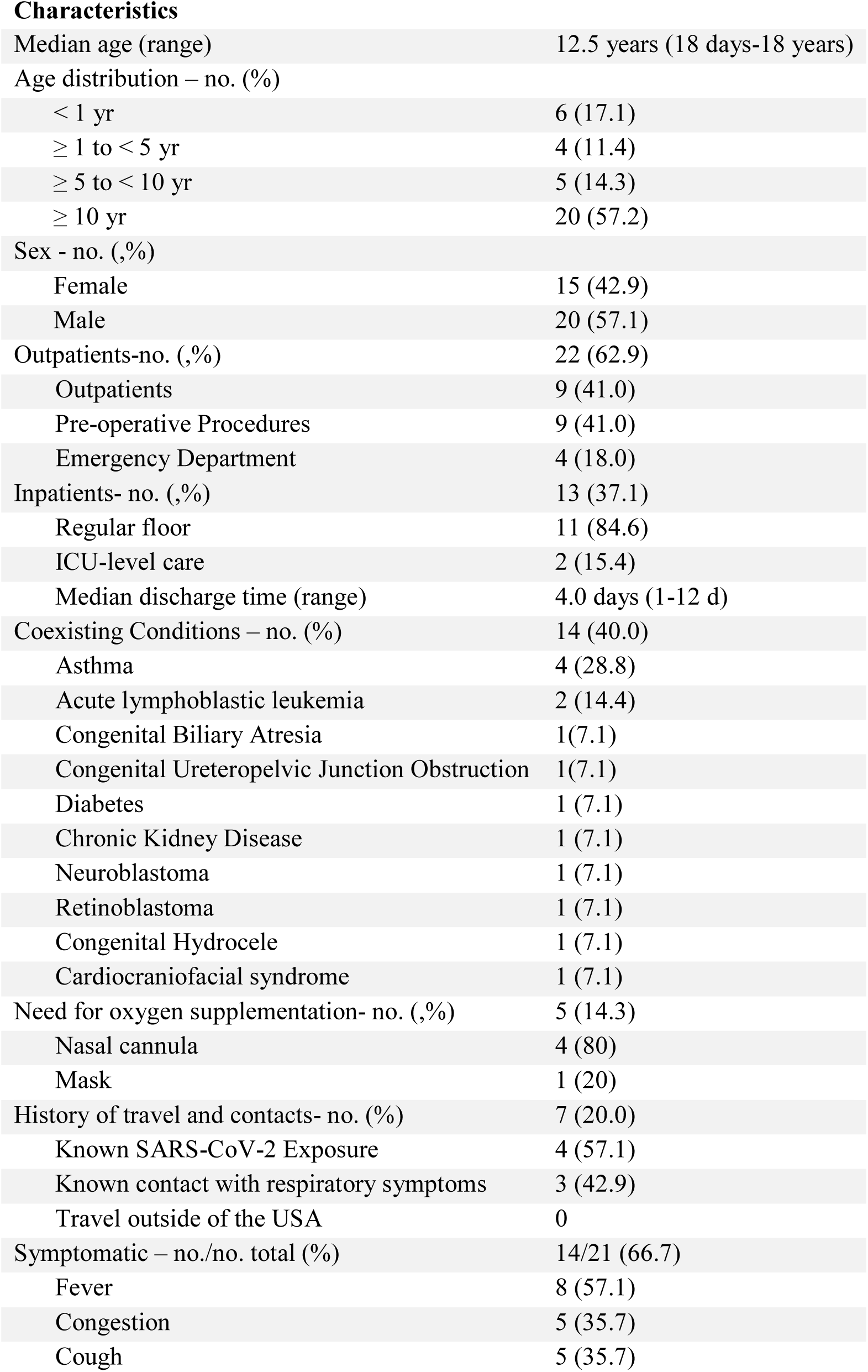
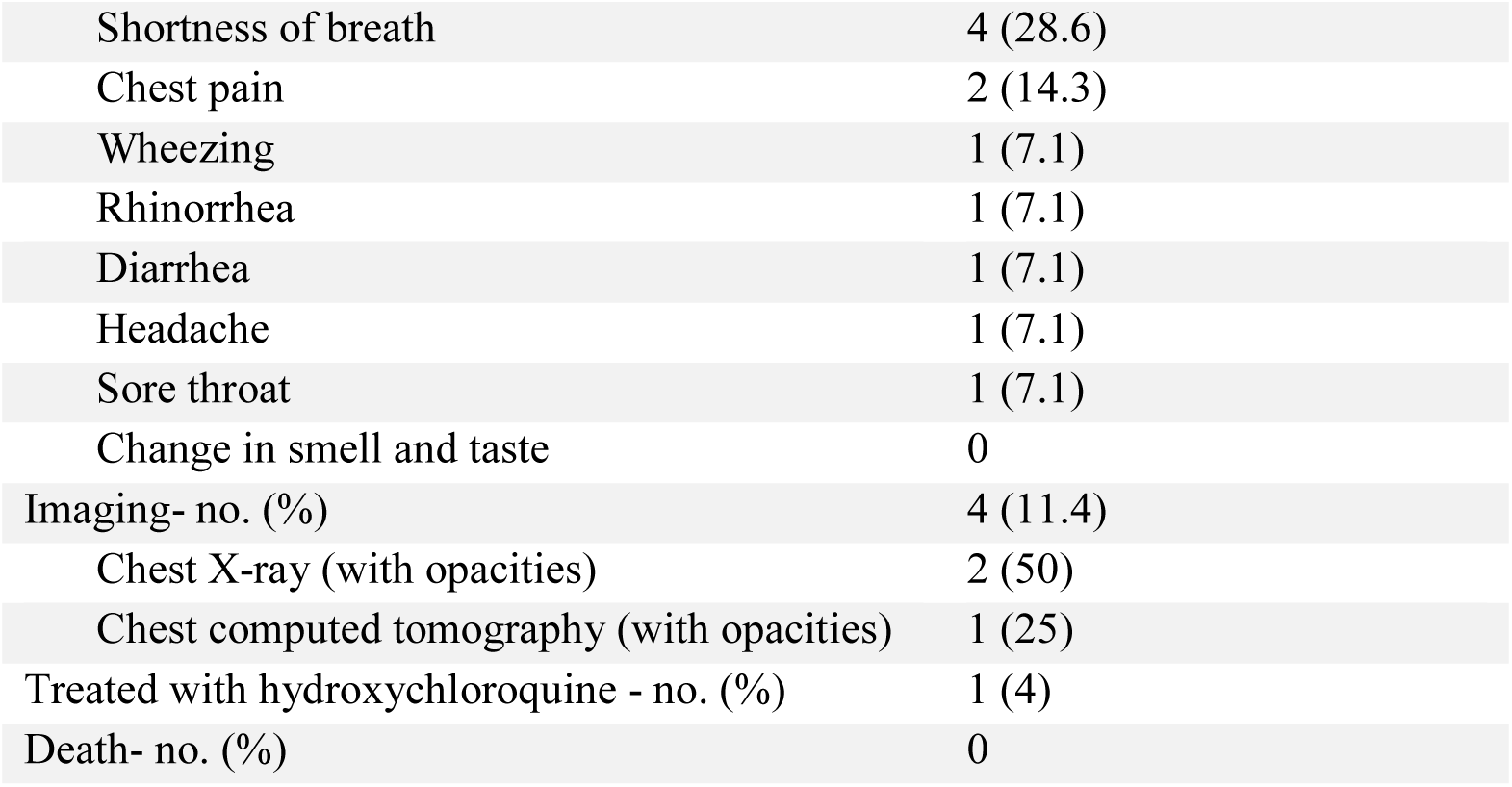
Epidemiologic Characteristics and Clinical Features of 35 Children Positive for SARS-CoV-2.

The median viral load obtained from all positive results was 1.6 × 10^6^ copies/mL (range: 2.7 × 10^2^ to 2.8 × 10^7^ copies/mL). The median SARS-CoV-2 viral load was higher in symptomatic than asymptomatic patients (2.4×10^7^ vs 1.2×10^4^ copies/mL, *p*=0.02). All patients <5 years old had higher viral loads (1.5×10^8^ vs 7.4 × 10^5^ copies/mL, *p*=0.04) and were symptomatic, corroborating the findings of prior studies demonstrating correlation between disease severity, viral load and younger age in children^8,9^. No difference in viral load was observed between those with chronic underlying conditions and those without (1.6×10^6^ vs 5.0 × 10^6^ copies/mL, *p*=0.3). Interestingly, one co-infection with human metapneumovirus was observed in a young infant^10^. Three of 5 patients with repeated testing were persistently positive for SARS-CoV-2 RNA for up to 16 days.

In our cohort, one child with standard risk B-cell acute lymphoblastic leukemia (ALL) was treated with a 4-day course of hydroxychloroquine. The patient was initially asymptomatic but developed symptoms 3 weeks into his hospital stay and was started on hydroxychloroquine with a plan for close cardiac monitoring. Within four days, his symptoms completely resolved, however, PCR continues to be positive 6 weeks after initial positive.

WGS of SARS-CoV-2 isolates from our 35 patients revealed 97 unique single-nucleotide variants (SNVs) and 7 insertions/deletions (IN/DELs) compared to the Wuhan isolate (NC-045512.2)^11^, with an average of 8.9 unique variations per isolate (0-14) (Fig.1a, Extended Data Table 1). These variants were located in the 5’UTR (n=1), pp1a (n=42), pp1ab (n=16), S (n=12), ORF3a (n=4), E (n=1), M (n=3), ORF6 (n=2), ORF7a (n=2), ORF8 (n=2), N (n=9) and stem-loop II of the 3’UTR (n=2) (Fig.1a, 1b). Of the 97 unique SNVs, 56 were non-synonymous, 30 were synonymous, and 3 were intergenic (Fig.1b). The predominance of non-synonymous variations across different ORFs has been previously documented, and highlights the evolution of SARS-CoV-2 during the course of the pandemic^12^. Five of 7 IN/DELs caused a frame-shift mutation in pp1a while 2 were present in the S protein. Examination of ORFs pp1a/ab and S with the highest number of SNVs for positive selection using Ka/Ks ratio, lacked statistical support (*p*≥0.05). Notably, the recently described D614G mutation in the S protein^13^, caused by nucleotide G-to-A substitution at position 23,403 in the Wuhan reference strain NC_045512.2 (Fig. 1a), was present in 33/35 (94.3%) isolates. This mutation was shown to be rapidly fixed in isolates from Europe and North America and has been postulated to play an important role in viral egress and enhancement of interaction between receptor-binding-domain of the S protein with viral entry receptor ACE2. Estimated evolutionary rate calculated using metadata from each isolate was 6.4 × 10^-4^ substitutions per site per year or 19.1 substitutions per year (Fig.1c, Extended Data Fig. 1). Our findings are concordant with the mutation rate of 6.0 × 10^-4^ substitutions per site per year reported by a recent study after analyzing 7,666 high-quality SARS-CoV-2 genomes from the GISAID database^14^. Remarkably, the mutation rate of SARS-CoV-2 is comparable to other RNA viruses, despite coronaviruses possessing the ability to encode a 3’-5’exoribonuclease -ExoN (nsp14) to proofread the complementary strand during genome replication, thus enhancing the fidelity of RNA-dependent RNA polymerase (RdRP) compared to other coronaviruses viruses^14–16^. The inferred time to most recent common ancestor (TMRCA) based on the molecular clock analysis of these isolates was 2019-12-04, which is comparable to the TMRCA of 2019-12-06, based on the analysis of 4,085 global isolates available in Nextstrain^17^, previously published data^14^, and with the start of the pandemic. Pairwise difference of just 8.9 variations per isolate between the Wuhan isolate and our isolates provides further support that these viruses share a recent common ancestor. Comparison of the 35 CHLA isolates to 966 SARS-CoV-2 genomes from the US and globally revealed that CHLA isolates clustered predominantly with other isolates from the US, but also with isolates from Europe and Australia (Fig.2). Two isolates from a sibling pair clustered together indicating familial transmission. This diversity among the CHLA isolates points to multiple potential introductions of the virus in Southern California from across the US and the world.

**Fig. 1.**
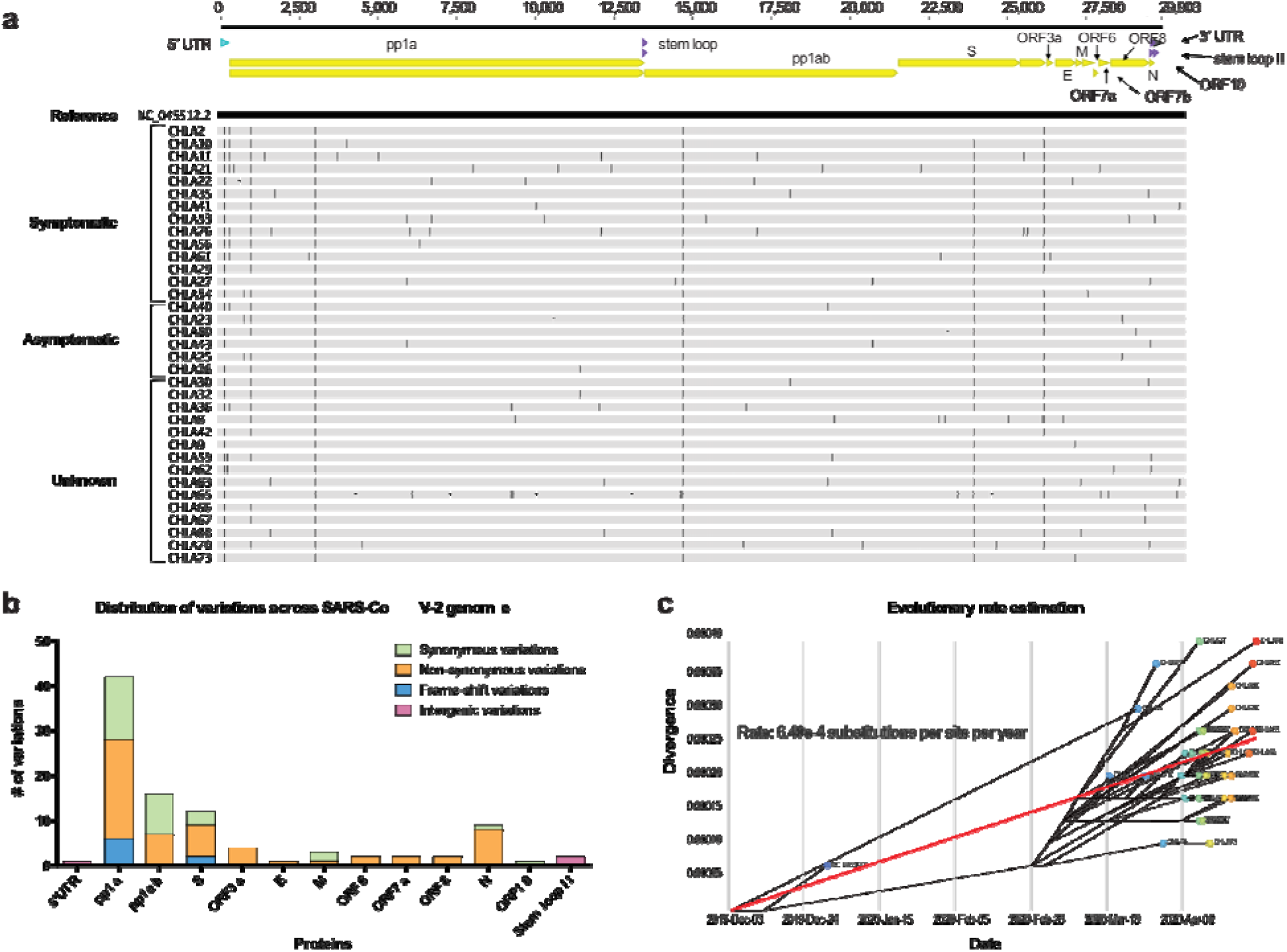
Single nucleotide variations (SNVs) and evolutionary rate of SARS-CoV-2 genomes obtained from 35 CHLA patients. **a**, Spread of SNVs across the genome in 35 CHLA isolates as compared to the Wuhan isolate (NC_045512.2). Vertical black lines on the genomes indicate presence of a SNV. Upper panel shows the structure of SARS-CoV-2 genome. **b**, Summary of characteristics of SNVs in open reading frames and non-coding regions. **c**, A time-resolved evolutionary rate estimation using 35 CHLA isolates.

**Fig. 2.**
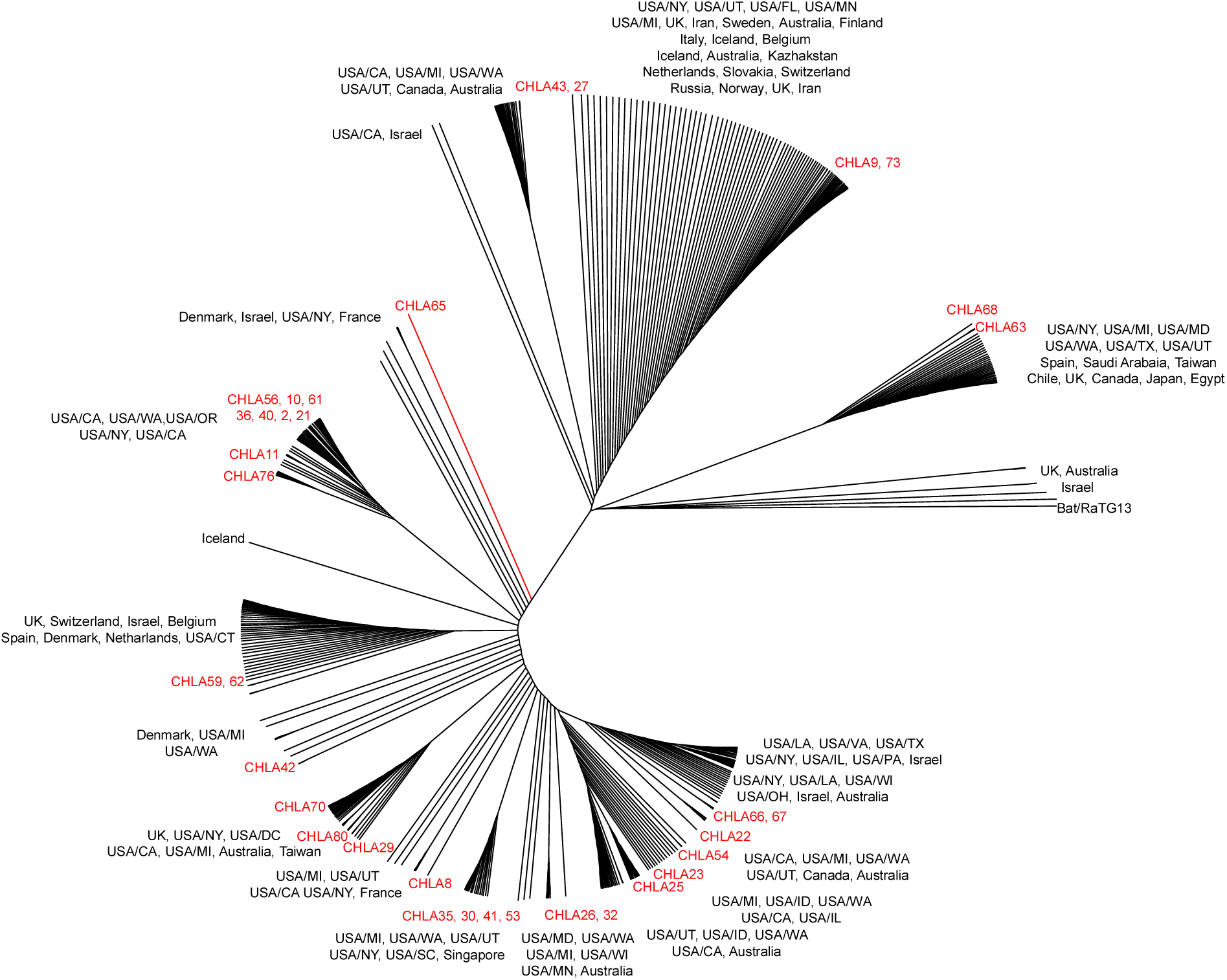
A whole-genome maximum likelihood phylogenetic tree showing genetic relatedness between SARS-CoV-2 isolates. Thirty-five isolates obtained from patients at CHLA and 931 previously sequenced isolates of different geographical origins from Global Initiative on Sharing All Influenza Data (GISAID) were used for phylogenetic investigation. CHLA isolate names and their branches are highlighted in red. Bat isolate is highlighted in blue. Isolates from the same family are labelled.

The idea of linking viral genetic diversity to disease severity is intriguing. Studies examining viral genomes during the Ebola 2013-2016 epidemic identified a single non-synonymous mutation in the viral glycoprotein, which increased its infectivity and severity in humans^4^. Whether genomic diversity in SARS-CoV-2 genome predicts disease severity remains to be determined. In our cohort, comparison of viral genomes did not identify variations solely present in symptomatic or asymptomatic patients (Fig. 1a). In fact, only 6 of 97 variations across the viral genome were present in more than 5 isolates regardless of the disease phenotype of the patient (Extended Data Table 1). Absence of shared variations determining disease phenotype points to host factors being the primary determinant of disease severity. Similar findings have been reported by a recent study of adult patients examining viral genomes from 112 patients^12^.

## Conclusion

Examination of clinical features in our cohort suggests that presentation of COVID-19 in children is multifaceted. We observed higher disease severity in younger children and disease manifestation correlated with viral load. Unlike previously suggested, the majority of our patients were symptomatic when tested, however, one-third were asymptomatic. These findings have direct implications for infection control within the hospital as it highlights the importance of screening patients before hospital admittance to investigate asymptomatic shedding and to avoid exposures. Sequencing of the viral genomes provided a glimpse into the viral genetic diversity in the circulating strains of SARS-CoV-2 in Southern California, which has thus far been lacking. We observed limited variations between the isolates. Nevertheless, the majority of these variations led to an amino-acid change in the viral protein, possibly indicating an on-going adaptation of the virus in human population. Most importantly, no variation was associated with disease manifestation. Our study presents the first pediatric cohort examining clinical, molecular, and epidemiological characteristics of pediatric COVID-19 infections in the US.

## Methods

### Study Design

We identified all positive patients tested at Children’s Hospital Los Angeles for detection of SARS-CoV-2 from nasopharyngeal swabs submitted between March 13 to May 11, 2020. A total of 35 patients were enrolled in this study. Demographic data including age, gender, location of admission, coexisting conditions, antimicrobial therapy, modes of oxygen supplementation, history of travel and contacts, clinical signs and symptoms (e.g. fever, congestion, cough, shortness of breath, wheezing, chest pain, rhinorrhea, diarrhea, headache, sore throat, and change in smell and taste), and radiographic findings (e.g. chest X-ray and chest computed tomography) were obtained from the electronic medical record.

### qRT–PCR assay for SARS-CoV-2 RNA

Nasopharyngeal swabs were sent to the Clinical Virology Laboratory at Children’s Hospital Los Angeles. Total nucleic acid was extracted from the samples using the NucliSENS easyMag (bioMerieux, France) and qRT-PCR was performed using the CDC 2019-Novel Coronavirus (2019-nCoV) Real-Time RT-PCR assay that has been granted emergency use authorization (EUA) by the U.S. Food and Drug Administration. A positive result for SARS-CoV-2 detection was determined by amplification of both N1 and N2 viral targets using a cut-off of Ct value < 40.

### Viral genome library construction and sequencing

WGS from extracted RNA was performed as previously described using Paragon Genomics CleanPlex SARS-CoV-2 Research and Surveillance NGS Panel^18^. Briefly, cDNA was synthesized by combining 11 μL of sample, with 3 μL of RT primer Mix DP and incubated for 5 minutes at 650C. 5 μL of RT Buffer DP and 1 μL of RT Enzyme were then added to the mix and incubated for 10 minutes at 80°C then for 80 minutes at 42°C. cDNA was then purified using 2.2X beads-to-sample ratio of CleanMag Magnetic Beads and 70% Ethanol. Separate multiplex-PCR reactions were then setup for primer pool 1 and 2 using 5 μL of purified cDNA, 2 μL of nuclease-free water, 2 μL of 5X mPCR Mix, and 1 μL of 10X SARS-CoV-2 Primer Pool 1/2. PCR conditions used were as follows: initial denaturation – 95°C for 10 minutes, 10 cycles of denaturation (98°C for 15 seconds) and annealing/extension (60°C for 5 minutes), hold at 100°C. Reactions for each primer pool were then combined and purification was performed using 1.3X beads-to-sample ratio of CleanMag Magnetic Beads and 70% Ethanol. Digestion reaction was then setup and incubated for 10 minutes at 37°C using 10 μL of purified PCR product, 7 μL of nuclease-free water, 2 μL of CP Digestion Buffer and 1 μL of CP Digestion Reagent. Digested libraries were then purified using 1.3X beads-to-sample ratio of CleanMag Magnetic Beads and 70% Ethanol. Second PCR reaction was then setup using 10 μL of purified libraries, 18 μL of nuclease-free water, 8 μL of 5X Second PCR Mix, and 2 μL each of i5 and i7 Dual-Indexed PCR Primer for Illumina. PCR conditions used were as follows: initial denaturation – 95°C for 10 minutes, 25 cycles of denaturation (98°C for 15 seconds) and annealing/extension (60°C for 75 seconds), hold at 10°C. 1X beads-to-sample ratio of CleanMag Magnetic Beads and 70% Ethanol was then used for purification to obtain the final library.

Libraries were quantified using the Agilent TapeStation High Sensitivity D1000 screen tape assay. Libraries were normalized to approximately 7nM, re-quantified and pooled to a final concentration of 4nM; pooled libraries were denatured and diluted according to Illumina protocols and loaded on the MiSeq at 10pM. Paired-end and dual-indexed 2x150bp sequencing was done using Micro Kit v2 (300 Cycles). Sample performance was selected based on the following metrics: average depth ≥ 1000x, percent bases covered at 10x ≥ 80%

### Consensus genome assembly

Nucleotide sequences were aligned with NovoAlign. Coverage profiles, variant calls and consensus genomes were generated using an in-house software system - LUBA. Consensus sequences were built by adjusting the reference genome at high allele frequency SNV and indel loci. Base quality adjusted pileup was generated, and the alternative bases and indels that accounted for more than 50% of the pileup were inserted into the reference sequence.

### Consensus genome comparison and phylogenetic analysis

Consensus genomes obtained for the 35 CHLA isolates was compared to the Wuhan isolate (NC_045512.2)^3^ using SARS-CoV-2 Genome App v1.1 (https://cov2annot.cpmbiodev.net) to identify synonymous, non-synonymous and intergenic variations.

For phylogenetic analysis, consensus sequence for each isolate was analyzed with the Virus Genome Tracker tool in CHLA COVID-19 Analysis Research Database (CARD)^2^. Variants were called, and the profile was compared against the global collection of about 30,000 available virus sequences in CARD. The closest isolates with identical or the most similar variant profiles were picked and classified by degrees of relatedness. Closest matches for all the 35 sequences were merged into a list of 969 unique isolates, which included 909 good quality (<500 missing bases) external isolates from the global database, 35 sequences included in this study, and additional 25 CHLA isolates not included in this study. Missing bases “N” were trimmed off from the 5’ and 3’ end, and the genomes were aligned to generate a multiple sequence alignment (MSA) with

MAFFT (version 7.460) using speed-oriented option - FFT-NS-i (iterative refinement method, two cycles) optimized for large datasets^19^. The resulting MSA was then manually checked and 3 external sequences were removed due to low-quality. The MSA was then trimmed by 65 bases from the 5’ end, and 66 bases from the 3’ end to remove low coverage regions. The final MSA had 966 sequences. Multiple copies of sequences that were 100% identical were then removed resulting in a final alignment of 589 sequences. MSA was then analyzed using MEGA-X ^20^ for evolutionary history inference using the Maximum Likelihood method and General Time Reversible model assuming protein-coding sequences. Positions with over 2% alignment gaps, missing data, and ambiguous bases were excluded. The bat coronavirus RaTG13/Yunnan/2013^11^ sequence was included in MSA for tree building as the outgroup to find the root of the tree. The phylogenetic tree was visualized in FigTree v1.4.4^21^.

### Evolutionary rate estimation

Rate estimation and visualization was performed using bioinformatics tools provided by Nextstrain^17^. MSA of the 35 CHLA isolates and the Wuhan isolate (NC_045512.2)^3^ was generated using MAFFT (version 7.453)^19^. A Maximum likelihood tree using Bayesian information criteria was generated with IQ-TREE (version 1.15.0)^22^ using GTR substitution model. The resulting rate estimation and phylogeny was then time-resolved using TreeTime^23^ and visualized using auspice^17^.

### Statistical analysis

Differences of Ct values were compared using Mann-Whitney test. P-value for selection was calculated using Fisher’s exact test for selection in MEGA.

### Ethics approval

Study design conducted at Children’s Hospital Los Angeles was approved by the Institutional Review Board under IRB CHLA-16-00429.

## Reporting summary

Further information on research design is available in the Nature Research Reporting Summary linked to this article.

## Data Availability

Whole genomes constructed for the purpose of this study will be submitted to GISAID and NCBI’s global SARS-CoV-2 genome database.

## Data Availability

The data shown in the manuscript are available upon request from the corresponding author. Nucleotide sequences of all 35 CHLA isolates have been submitted to NCBI and GISAID.

## Acknowledgments

We would like to acknowledge the staff members of Virology laboratory and Center for Personalized Medicine at Children’s Hospital Los Angeles for their dedication towards providing excellent patient care. We would also like to acknowledge the frontline healthcare workers at our institution and across the world who remain devoted in the fight against COVID-19. We would also like to acknowledge NCBI, GISAID, and Nextstrain for providing valuable resources for SARS-CoV-2 genomics.

## Author Contributions

U.P., R.Y., and J.D.B contributed to the study design. U.P., R.Y., J.M. B, and J.D.B contributed to the collection of clinical specimens, acquisition, analysis, and interpretation of the clinical data. U.P, D.O, performed library construction and whole genome sequencing. U.P., M.B., L.S., A.R., D.T.M, and X.G. contributed to analysis of genomic sequences. U.P and R.Y performed the statistical analysis. U.P, R.Y, M.P., M.B., A.R., L.S., D.T.M., O.D., J.A.B., A.R.J., J.M.B., X.G., J.D.B contributed to writing the manuscript. All authors approved the manuscript.

## Competing interests

The authors declare no competing interest

---

**Extended Data Fig. 1.**
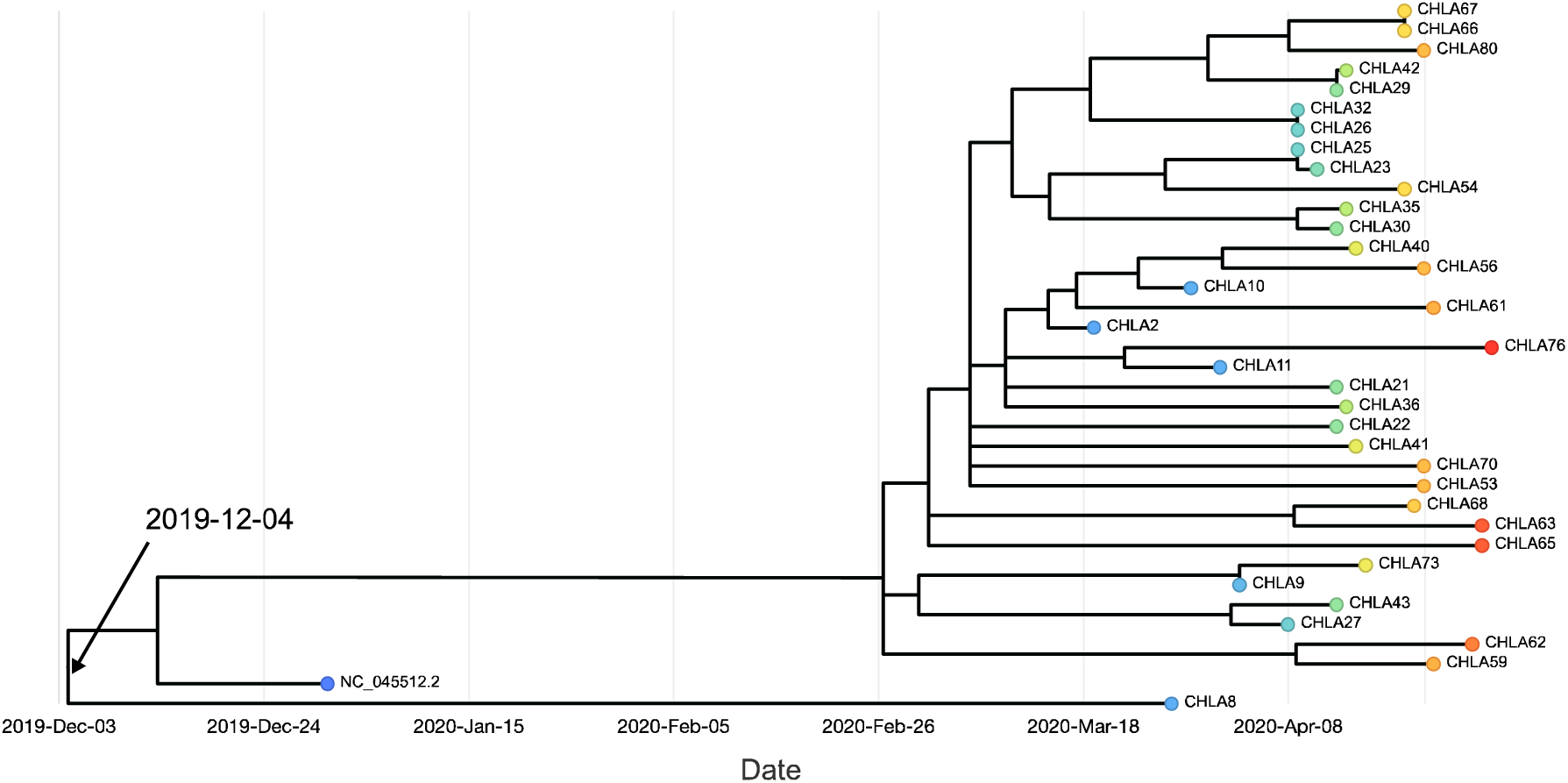
A time-resolved Maximum likelihood phylogenetic tree of 35 CHLA isolates. Each sample is represented by a single color. Collection date for each sample is shown in the x-axis. Time to most-recent ancestor per analysis is shown.

**Extended Data Table 1.**
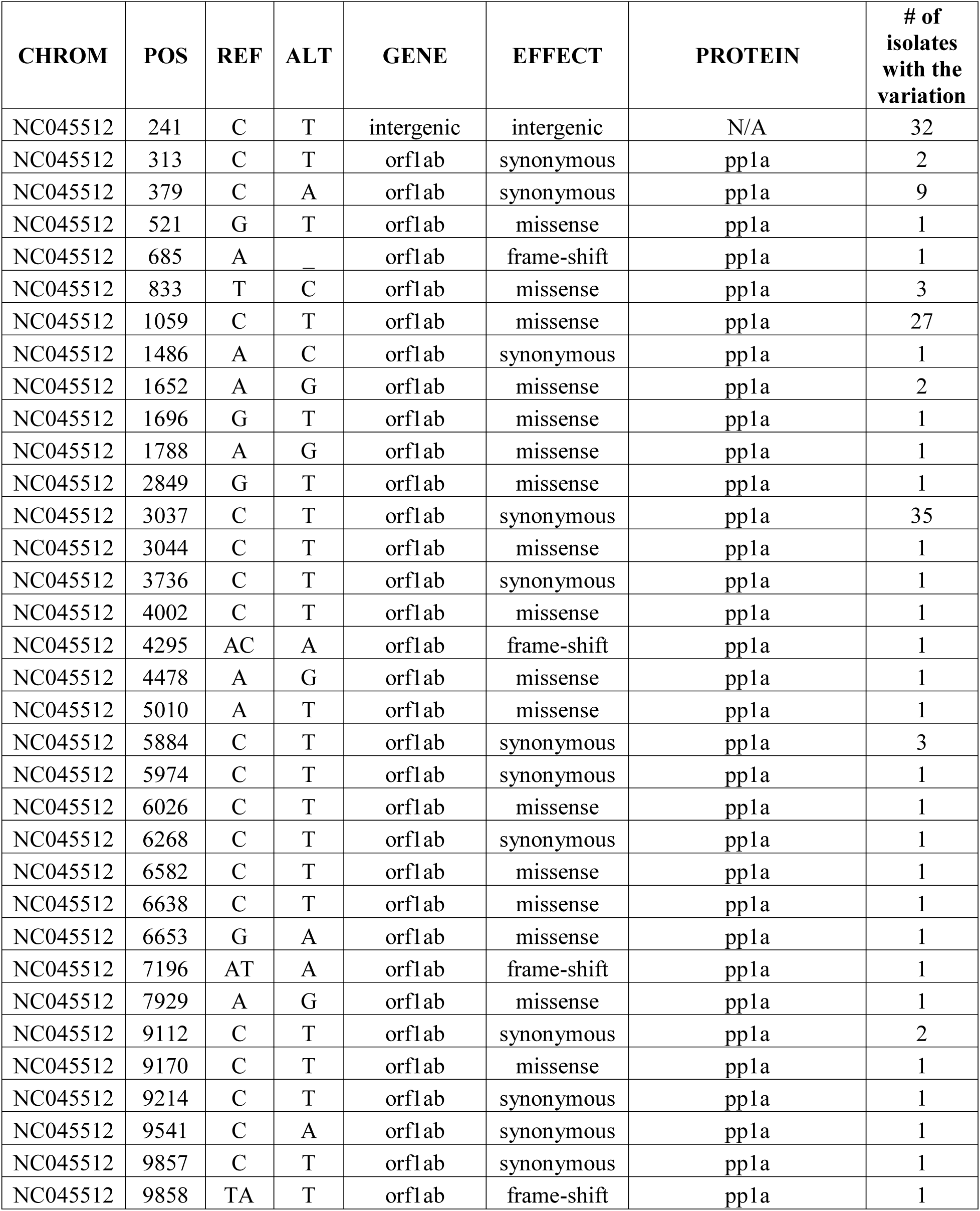
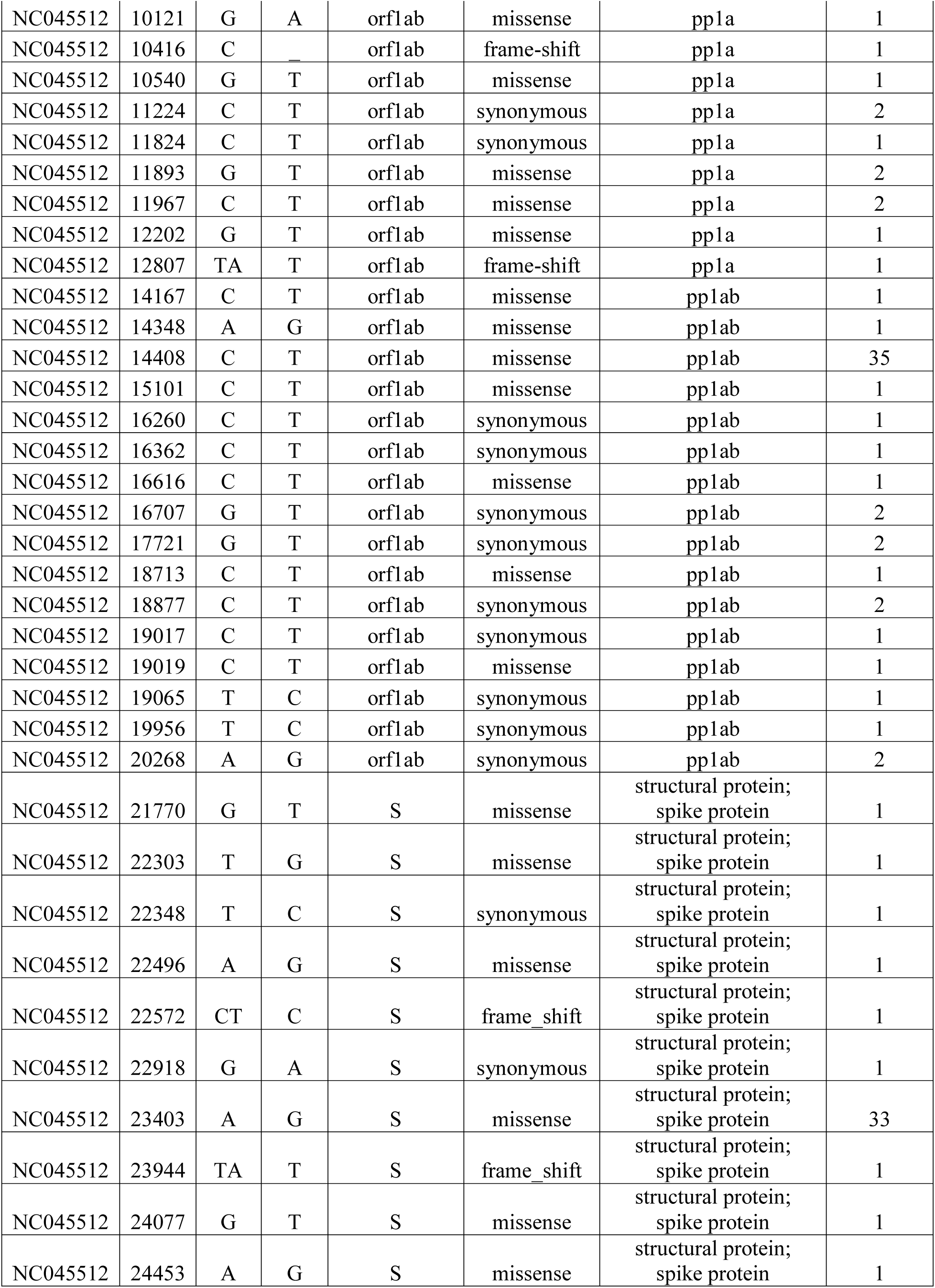
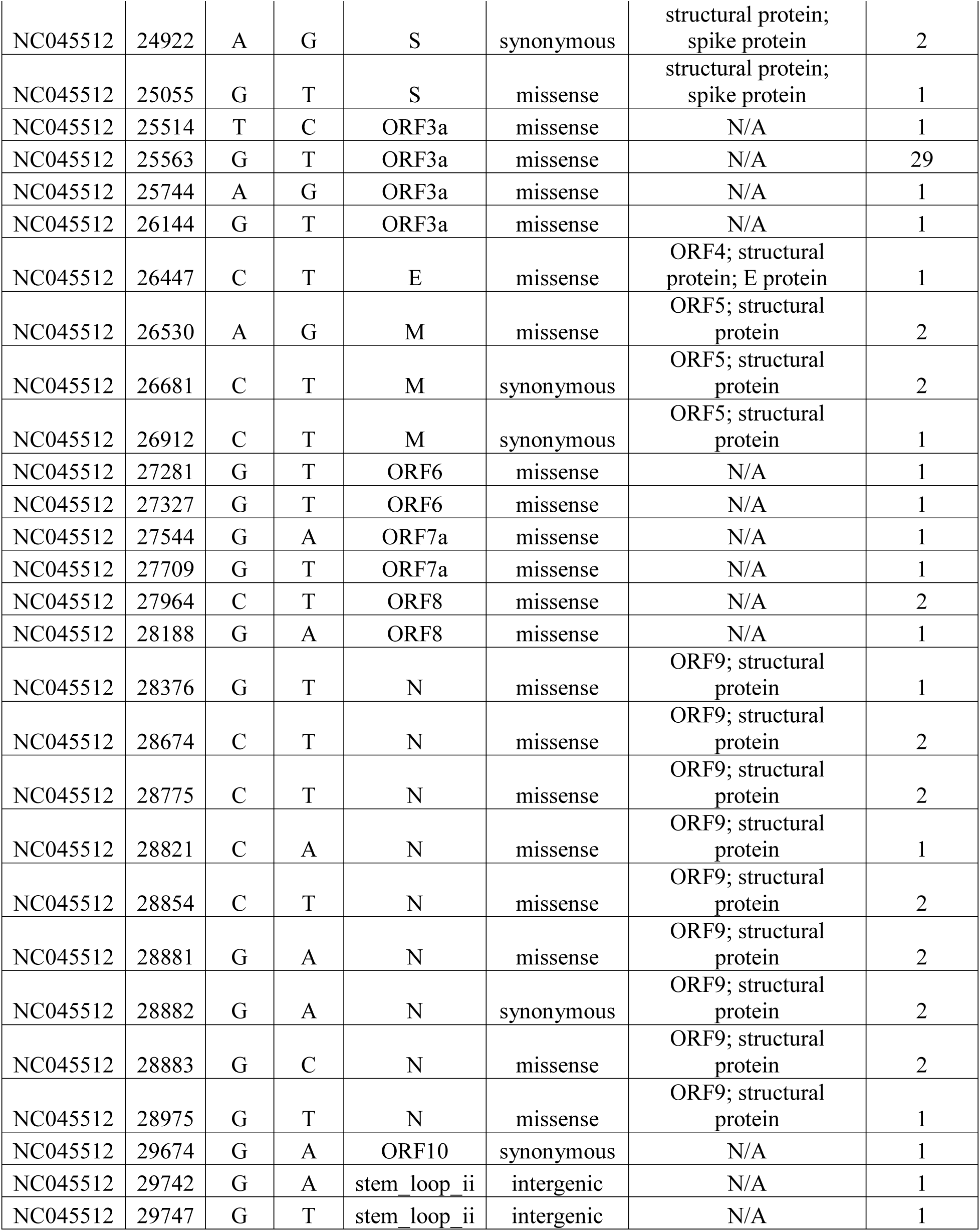
Position, effect and isolate count for variations identified across the viral genome for 35 CHLA isolates.

| CHROM | POS | REF | ALT | GENE | EFFECT | PROTEIN | # of isolates with the variation |
| --- | --- | --- | --- | --- | --- | --- | --- |
| NC045512 | 241 | C | T | intergenic | intergenic | N/A | 32 |
| NC045512 | 313 | C | T | orf1ab | synonymous | pp1a | 2 |
| NC045512 | 379 | C | A | orf1ab | synonymous | pp1a | 9 |
| NC045512 | 521 | G | T | orf1ab | missense | pp1a | 1 |
| NC045512 | 685 | A | _ | orf1ab | frame-shift | pp1a | 1 |
| NC045512 | 833 | T | C | orf1ab | missense | pp1a | 3 |
| NC045512 | 1059 | C | T | orf1ab | missense | pp1a | 27 |
| NC045512 | 1486 | A | C | orf1ab | synonymous | pp1a | 1 |
| NC045512 | 1652 | A | G | orf1ab | missense | pp1a | 2 |
| NC045512 | 1696 | G | T | orf1ab | missense | pp1a | 1 |
| NC045512 | 1788 | A | G | orf1ab | missense | pp1a | 1 |
| NC045512 | 2849 | G | T | orf1ab | missense | pp1a | 1 |
| NC045512 | 3037 | C | T | orf1ab | synonymous | pp1a | 35 |
| NC045512 | 3044 | C | T | orf1ab | missense | pp1a | 1 |
| NC045512 | 3736 | C | T | orf1ab | synonymous | pp1a | 1 |
| NC045512 | 4002 | C | T | orf1ab | missense | pp1a | 1 |
| NC045512 | 4295 | AC | A | orf1ab | frame-shift | pp1a | 1 |
| NC045512 | 4478 | A | G | orf1ab | missense | pp1a | 1 |
| NC045512 | 5010 | A | T | orf1ab | missense | pp1a | 1 |
| NC045512 | 5884 | C | T | orf1ab | synonymous | pp1a | 3 |
| NC045512 | 5974 | C | T | orf1ab | synonymous | pp1a | 1 |
| NC045512 | 6026 | C | T | orf1ab | missense | pp1a | 1 |
| NC045512 | 6268 | C | T | orf1ab | synonymous | pp1a | 1 |
| NC045512 | 6582 | C | T | orf1ab | missense | pp1a | 1 |
| NC045512 | 6638 | C | T | orf1ab | missense | pp1a | 1 |
| NC045512 | 6653 | G | A | orf1ab | missense | pp1a | 1 |
| NC045512 | 7196 | AT | A | orf1ab | frame-shift | pp1a | 1 |
| NC045512 | 7929 | A | G | orf1ab | missense | pp1a | 1 |
| NC045512 | 9112 | C | T | orf1ab | synonymous | pp1a | 2 |
| NC045512 | 9170 | C | T | orf1ab | missense | pp1a | 1 |
| NC045512 | 9214 | C | T | orf1ab | synonymous | pp1a | 1 |
| NC045512 | 9541 | C | A | orf1ab | synonymous | pp1a | 1 |
| NC045512 | 9857 | C | T | orf1ab | synonymous | pp1a | 1 |
| NC045512 | 9858 | TA | T | orf1ab | frame-shift | pp1a | 1 |
| NC045512 | 10121 | G | A | orf1ab | missense | pp1a | 1 |
| NC045512 | 10416 | C | _ | orf1ab | frame-shift | pp1a | 1 |
| NC045512 | 10540 | G | T | orf1ab | missense | pp1a | 1 |
| NC045512 | 11224 | C | T | orf1ab | synonymous | pp1a | 2 |
| NC045512 | 11824 | C | T | orf1ab | synonymous | pp1a | 1 |
| NC045512 | 11893 | G | T | orf1ab | missense | pp1a | 2 |
| NC045512 | 11967 | C | T | orf1ab | missense | pp1a | 2 |
| NC045512 | 12202 | G | T | orf1ab | missense | pp1a | 1 |
| NC045512 | 12807 | TA | T | orf1ab | frame-shift | pp1a | 1 |
| NC045512 | 14167 | C | T | orf1ab | missense | pp1ab | 1 |
| NC045512 | 14348 | A | G | orf1ab | missense | pp1ab | 1 |
| NC045512 | 14408 | C | T | orf1ab | missense | pp1ab | 35 |
| NC045512 | 15101 | C | T | orf1ab | missense | pp1ab | 1 |
| NC045512 | 16260 | C | T | orf1ab | synonymous | pp1ab | 1 |
| NC045512 | 16362 | C | T | orf1ab | synonymous | pp1ab | 1 |
| NC045512 | 16616 | C | T | orf1ab | missense | pp1ab | 1 |
| NC045512 | 16707 | G | T | orf1ab | synonymous | pp1ab | 2 |
| NC045512 | 17721 | G | T | orf1ab | synonymous | pp1ab | 2 |
| NC045512 | 18713 | C | T | orf1ab | missense | pp1ab | 1 |
| NC045512 | 18877 | C | T | orf1ab | synonymous | pp1ab | 2 |
| NC045512 | 19017 | C | T | orf1ab | synonymous | pp1ab | 1 |
| NC045512 | 19019 | C | T | orf1ab | missense | pp1ab | 1 |
| NC045512 | 19065 | T | C | orf1ab | synonymous | pp1ab | 1 |
| NC045512 | 19956 | T | C | orf1ab | synonymous | pp1ab | 1 |
| NC045512 | 20268 | A | G | orf1ab | synonymous | pp1ab | 2 |
| NC045512 | 21770 | G | T | S | missense | structural protein;<br>spike protein | 1 |
| NC045512 | 22303 | T | G | S | missense | structural protein;<br>spike protein | 1 |
| NC045512 | 22348 | T | C | S | synonymous | structural protein;<br>spike protein | 1 |
| NC045512 | 22496 | A | G | S | missense | structural protein;<br>spike protein | 1 |
| NC045512 | 22572 | CT | C | S | frame_shift | structural protein;<br>spike protein | 1 |
| NC045512 | 22918 | G | A | S | synonymous | structural protein;<br>spike protein | 1 |
| NC045512 | 23403 | A | G | S | missense | structural protein;<br>spike protein | 33 |
| NC045512 | 23944 | TA | T | S | frame_shift | structural protein;<br>spike protein | 1 |
| NC045512 | 24077 | G | T | S | missense | structural protein;<br>spike protein | 1 |
| NC045512 | 24453 | A | G | S | missense | structural protein;<br>spike protein | 1 |
| NC045512 | 24922 | A | G | S | synonymous | structural protein;<br>spike protein | 2 |
| NC045512 | 25055 | G | T | S | missense | structural protein;<br>spike protein | 1 |
| NC045512 | 25514 | T | C | ORF3a | missense | N/A | 1 |
| NC045512 | 25563 | G | T | ORF3a | missense | N/A | 29 |
| NC045512 | 25744 | A | G | ORF3a | missense | N/A | 1 |
| NC045512 | 26144 | G | T | ORF3a | missense | N/A | 1 |
| NC045512 | 26447 | C | T | E | missense | ORF4; structural<br>protein; E protein | 1 |
| NC045512 | 26530 | A | G | M | missense | ORF5; structural<br>protein | 2 |
| NC045512 | 26681 | C | T | M | synonymous | ORF5; structural<br>protein | 2 |
| NC045512 | 26912 | C | T | M | synonymous | ORF5; structural<br>protein | 1 |
| NC045512 | 27281 | G | T | ORF6 | missense | N/A | 1 |
| NC045512 | 27327 | G | T | ORF6 | missense | N/A | 1 |
| NC045512 | 27544 | G | A | ORF7a | missense | N/A | 1 |
| NC045512 | 27709 | G | T | ORF7a | missense | N/A | 1 |
| NC045512 | 27964 | C | T | ORF8 | missense | N/A | 2 |
| NC045512 | 28188 | G | A | ORF8 | missense | N/A | 1 |
| NC045512 | 28376 | G | T | N | missense | ORF9; structural<br>protein | 1 |
| NC045512 | 28674 | C | T | N | missense | ORF9; structural<br>protein | 2 |
| NC045512 | 28775 | C | T | N | missense | ORF9; structural<br>protein | 2 |
| NC045512 | 28821 | C | A | N | missense | ORF9; structural<br>protein | 1 |
| NC045512 | 28854 | C | T | N | missense | ORF9; structural<br>protein | 2 |
| NC045512 | 28881 | G | A | N | missense | ORF9; structural<br>protein | 2 |
| NC045512 | 28882 | G | A | N | synonymous | ORF9; structural<br>protein | 2 |
| NC045512 | 28883 | G | C | N | missense | ORF9; structural<br>protein | 2 |
| NC045512 | 28975 | G | T | N | missense | ORF9; structural<br>protein | 1 |
| NC045512 | 29674 | G | A | ORF10 | synonymous | N/A | 1 |
| NC045512 | 29742 | G | A | stem_loop_ii | intergenic | N/A | 1 |
| NC045512 | 29747 | G | T | stem_loop_ii | intergenic | N/A | 1 |

## Notes

### Competing Interest Statement

The authors have declared no competing interest.

### Funding Statement

No external funding was received for this project.

### Author Declarations

The study was conducted at CHLA, a freestanding tertiary care pediatric medical center, and was approved by the CHLA Institutional Review Board.

